# A qualitative study investigating the acceptability of cervical screening and self-sampling in postnatal women at 6-week postnatal check-up

**DOI:** 10.64898/2025.12.09.25341885

**Authors:** Rebecca Newhouse, Lorna McWilliams, Holly Baker-Rand, Victoria Cullimore, Emma Crosbie, Sudha Sundar, Jo Morrison

**Author notes:** Corresponding author – Jo Morrison Email –. joint first authors (Rebecca Newhouse, Lorna McWilliams).

## Abstract

**Introduction:** There is a lack of evidence to support clinical recommendations to delay cervical screening to 12-weeks postnatal. In previous studies, half of women were out of date for screening by the end of pregnancy and the majority would be more likely to take up cervical screening, if offered at the 6-week postnatal check-up. We explored views about postnatal cervical screening the acceptability of offering cervical screening, using conventional and urine self-sampling, earlier within the postnatal period.

**Methods:** A cross-sectional qualitative design was used with recruitment from a larger questionnaire-based study. Twenty-six online semi-structured interviews were conducted with 26 pregnant or recently pregnant participants. Interviews were transcribed pseudonymised. A topic guide was developed, and data analysed using inductive reflexive thematic analysis.

**Results:** Three themes were generated from qualitative analysis of verbatim interview transcripts: 1) A window of opportunity; 2) Am I ready yet? Postpartum recovery; and 3) Neglect of women’s health in and around pregnancy. Overall, there was a perception that women’s health was not a priority in the postnatal period compared with their babies.

**Conclusion:** This is the first study to use qualitative interview methods to explore women’s views about the offer of cervical screening alongside the postnatal check-up. Results support the feasibility of a clinical trial to test the accuracy and effect on uptake of offering cervical screening at the postnatal check-up, although recognised it might be too soon for some. This should be considered in future feasibility research which includes assessment of concurrent acceptability.

**Patient or Public Contribution:** This study was performed following focus groups in a quality improvement project, designed to increase uptake of cervical screening in women and people who were pregnant or recently pregnant. The suggestion for combining cervical screening with the routine 6-week postnatal follow up was an idea generated by new parents and GP practice staff. The Somerset Maternity Voices group provided feedback on study materials, including the consent form and posters.

The semi-structured interview topic guide was designed following free-text comments in the pre-PINCS web-based survey, results of which are published separately. Female pregnant and recently pregnant people, regardless of current gender identity, were included in this study. In line with the Royal College of Obstetricians and Gynaecologists language guide, we will use ‘women’ to describe participants.

**Practitioner Points:**

- Women recognise the importance of cervical screening, with an increased awareness of need of self-care for the benefit of their child,
- There are increased practical barriers to attending screening as new parents and efforts to reduce barriers would likely improve uptake.
- Women felt frustration around receiving mixed messaging and disinformation around cervical screening in and around pregnancy and would welcome a shift in focus to maternal care aspects at the postnatal check-up.

## Introduction

The National Health Service Cervical Screening Programme (NHS CSP) call-recall system was introduced in the UK in 1988. Since its introduction, deaths from cervical cancer have reduced by 70% [1]. However, performance of the programme depends on coverage rates, with a target of 80% coverage [2]. At 67%, coverage rates in England in the under 50s were at an all-time low by 2022/23, and lower than 40% in some areas [3–5]. Sadly, coverage is especially poor in higher-risk groups including those from areas of higher index of multiple deprivation (IMD), ethnic minorities, younger people with a cervix, and those with children aged less than 5 years [4,6,7].

Our previous quality improvement (QI) programme was driven by the diagnosis of cervical cancer in several young women, either during or shortly after a pregnancy, where diagnosis was made following symptoms, rather than screen-detected [8]. Most had locally advanced disease, requiring radical treatment, and sadly some went on to die from their disease, leaving young children without a mother. In all cases, opportunities for cervical screening, either during or shortly after a pregnancy had been missed. We found that half of new mothers were out of date with screening by the end of pregnancy and only half of these had participated in cervical screening by 6-months postnatal [8]. Simple measures, including targeted education of pregnant women and midwifery staff about cervical screening in pregnancy, improved cervical screening rates by 8%. This work included focus groups and canvassing of new mums, young women with a diagnosis of cervical cancer and primary care staff. One key idea for change, suggested by both women and primary care teams at a practice located in an area of higher socioeconomic deprivation, was to move postnatal cervical screening to coincide with the 6-week postnatal mother and baby check-up. Both groups suggested this would make attending easier for women, reducing barriers to attendance [9]. Self-sampling was also suggested to improve uptake for busy new mums, as this is acceptable to women [10], and is safe to use within a screening programme [11,12]. However, clinical trials have not specifically targeted this sizable under-screened population.

Current NHS CSP guidance, for those on routine recall (most recent screening test normal) whose screening becomes due in pregnancy, is that cervical screening should be delayed until 12-weeks postnatal [13]. All others should be offered screening during pregnancy, although we found that this is often misinterpreted, due to the commonly held belief that cervical screening didn’t need to, or shouldn’t, be performed in pregnancy [8]. There is limited evidence to support this recommendation to delay to 12-weeks with modern high-risk human papilloma virus (hrHPV)and liquid based cytology (LBC) cervical screening techniques. A randomised comparison of women with Papanicolaou smears, taken at 4, 6 and 8 weeks postnatal, demonstrated increased inflammatory changes at 4 weeks, although little difference between 6- and 8-week sampling [14]. This study pre-dates the introduction of LBC and HPV testing, which largely remove issues cause by inflammatory samples [15–18].

Recent work has focussed on self-sampling to improve uptake in those who have not been willing or able to take up offers of cervical screening [19–21]. However, no study has specifically evaluated the postnatal or pregnant cohort, who have low screening uptake rates [6,8]. A systematic review of self-sampling found that HPV self-sampling was generally highly acceptable [22], but there was concern about being able to reliably collect a self-sample in some who preferred clinician-collected samples. In this systematic review, women preferred the idea of home-based self-sampling to self-sampling at a clinic. Interestingly, in a pragmatic clinical trial of offering self-sampling to those out of date for screening, YouScreen, return rates of vaginal swabs was substantially lower in those sent kits to their home in the post compared with those offered at an opportunistic visit to primary care [20]. These and other data suggest that direct requests by health-care professionals during a consultation, and reducing as many practical barriers as possible, helps to improve cervical screening uptake [23]. Additionally, the survey of acceptability of self-sampling found that, although most would accept self-sampling in the future this was not universal [19]. Women preferred the idea of urine self-sampling (although this was only considered hypothetically), more so for those with minority ethnic backgrounds.

A qualitative systematic review of general practitioners (GPs) found that they viewed the postnatal check-up as an opportunity for relationship building and health promotion [24], although organisational barriers impacted motivation to provide the best care. A study looking at expectation of the those attending the postnatal check-up found that new parents appreciated this appointment, with it regarded as an ‘important milestone’ [25]. It was especially appreciated if the appointment combined mother and baby checks.

These findings support our hypothesis that providing an opportunity for cervical screening at an in-person postnatal check-up, would improve cervical screening uptake and aid more general health promotion for women, not just their babies. However, we wanted to explore this to understand in more depth maternal/birth parental attitudes to cervical screening, views on timing and methods of screening after birth, and the postnatal check-up, prior to embarking on a clinical trial to test this hypothesis. This in in line with the Medical Research Council guidance for evaluating complex interventions [26].

## Materials & methods

### Positionality

No one comes to research without preconceptions formed by their prior experiences. It is therefore important to be aware of these in the context of qualitative analysis. More than half of the authors have given birth themselves and are parents and at least one has had treatment for cervical abnormalities. All identify as White females and all but one have worked for over 4 years as obstetricians, have witnessed the effects physical and psychological birth trauma, and have spent many hours suturing women after vaginal births. These 6 authors have also seen women for follow up with vaginal examination at around 6-weeks postnatal to check healing and function. None of the authors have worked in primary care. Three had experience of qualitative methodology prior to this study and those who had not came with an open mind and eagerness to listen and learn. All the researchers are aware of the evidence of the effectiveness of cervical screening and support the cervical screening programme. RN, HBR, SS, EC and JM are colposcopists. LMcW is a female research fellow in psychology who has never given birth. She is interested in research which improves equity in cancer early detection including women’s health so found this an important topic to gather women’s perspectives on the intersection of cervical cancer screening and postnatal healthcare as it is largely neglected from the literature. JM is female gynaecological oncologist and researcher. She is a member of the NHS Cervical Screening Research, Innovation and Development Committee (2021-) and has personal experience of abnormal cervical screening and treatment. She has looked after several women diagnosed with cervical cancer during and shortly after pregnancy, all of whom had significant impact on healthcare workers as ‘second victims’. This work was precipitated by the death from cervical of a woman with young children, who had missed screening opportunities during and after pregnancies and is dedicated to her memory and her family.

### Design

A cross-sectional qualitative design was employed using semi-structured interviews. A critical realist perspective underpinned this study whereby the researchers believe that participant views expressed in this qualitative study are mediated by their social and cultural contexts [27].

### Sampling and participants

Participants in an anonymous web-based questionnaire were asked if they would be willing to participate in an hour-long interview to explore their views about postnatal cervical screening in more depth. Findings from the questionnaire study are presented elsewhere [28]. Study eligibility criteria were: female with a cervix, aged between 24.5 and 65 years, pregnant or had a pregnancy within 5-years. Women additionally had to be within 6-months of pregnancy to take part in the interview. 123/454 participants volunteered contact details, 40 of whom were beyond 6-months postnatal during the recruitment period in the present study. We had intended to purposely sample the volunteers, to include a range of participants, based on age, ethnicity, cervical screening history, and willingness to partake in a clinical trial of 6-week postnatal cervical screening with clinician taken samples and/or urine self-sampling. However, after approaching all 83 volunteers once by email, we included all 26 who agreed to participate.

### Data collection

Interviews were offered through Microsoft Teams or via telephone. Interviews lasted 16-45 minutes (median 25.5 minutes) and were conducted by one researcher (RN), a female obstetrics and gynaecology speciality trainee. A topic guide (Supplementary materials 1) was developed, informed by the participant responses to the questionnaire, to be used flexibly during interviews to explore how participants experienced cervical screening (or not), their views on cervical screening offered at the 6-week post-natal check-up and self-sampling.

Participants were also asked to elaborate on their views around taking part in a clinical research study about cervical screening at the 6-week post-natal check-up timepoint, based on their web-based questionnaire answers. They gave consent for the demographic information they reported in the questionnaire to be used in this sub-study: age, education attainment, employment status, number of children, birth of last child, ethnic group and when they had their last cervical screen (never, over 3-years ago or within the past 3-years). The interviewer used the questionnaire answers as prompts, where appropriate, throughout the interview. She was mindful about not imposing views or correcting misassumptions about screening. Data collection continued until the research team agreed that the data collected was sufficient to answer the study research question [29]. Interviews were audio-recorded, pseudonymised by removal of personal details and participants assigned a participant number for identification.

Recordings were sent for verbatim professional transcription. Audio recordings were retained to check for accuracy of transcription (RN) and then destroyed. Transcripts were checked for accuracy and any additional potentially identifying details within the transcripts were removed prior to analysis.

### Data analysis

Qualitative data were analysed using inductive reflexive thematic analysis to identify patterns in the data across the participants whilst acknowledging the role of the researchers in this process [30]. Data analysis was led by the lead author (RN) using Nvivo14 to code the data, with supervision from LMcW and input from JM throughout. Data were primarily coded at the manifest level, based on the language used by participants. RN and LMcW both coded two transcripts to discuss early coding as part of developing reflexive practice during analysis. Codes were developed iteratively and linked together using an online sticky note website to create an initial thematic map. Data extracts to illustrate initial themes were evaluated across the dataset with attention paid to negative cases to ensure these were also represented. The final thematic structure was deemed representative of the dataset based on several iterations of the written results.

Post-hoc exploratory analysis of quantitative data comparing the interview sub-study participants to the non-participants of the questionnaire study, to put comments of sub-study volunteers in context with the overall cohort, were evaluated for independence between categorical variables using Chi-square or Fisher’s exact tests. Statistical analysis was performed with Microsoft Excel [31] and GraphPad Prism software.[32]

## Results

Twenty-six pregnant or recently pregnant (within 6 months) participants were taken from the total questionnaire cohort of 454 [28]. Compared to the total cohort, interview volunteers were not significantly different in terms of age (median 30-34 years; χ^2^ (4) = 9.378; p = 0.052), ethnicity (χ^2^ (1) = 0.05977; p = 0.8069), employment status (χ^2^ (3) = 3.475; p = 0.324) or educational attainment (χ^2^ (2) = 2.50; p = 0.287) (see Table 1). Median parity was 1 in the interview group, compared to 2 in the non-interview group, although not significantly different (χ^2^ (3) = 4.811; p = 0.186). Most women had at least 1 child; two were pregnant when they provided demographic information prior to the interview. All but one participant reported regular engagement with cervical screening, which contextualises our findings. One had not had screening previously as they were too young prior to their pregnancy. However, 10/26 (38%) were out of date for screening, which was higher than the non-interview cohort (χ^2^ (2) = 10.28; p = 0.0059). This was presumably as this subgroup were more likely to be within 12 months of their last pregnancy, due to the requirement to have been within 6 months of pregnancy to volunteer for the interview sub-study (χ^2^ (4) = 24.85; p<0.0001). Individual participant numbers and characteristics are given in Table 2.

**Table 1.** Demographic data. Demographic data of the 26 participants in the interview study compared with non-participants from the original questionnaire cohort (total cohort n = 454)

|  | Interview cohort | Non-interview cohort | Inter-cohort comparison |
| --- | --- | --- | --- |
|  | **n = 26 (%)** | **n = 428 (%)** |  |
| **Age** |  |  | **p = 0.0523** |
| 24-29 years old | 10 (38%) | 72 (17%) |  |
| 30-34 years old | 11 (42%) | 180 (42%) |  |
| 35-39 years old | 4 (15%) | 131 (31%) |  |
| 40+ years old | 1 (4%) | 45 (11%) |  |
| **Number of children** |  |  | **p = 0.1862** |
| 0 | 1 (4%) | 27 (6%) |  |
| 1 | 15 (58%) | 162 (38%) |  |
| 2 | 6 (23%) | 178 (42%) |  |
| 3 or more | 4 (15%) | 61 (14%) |  |
| **Education** |  |  | **p = 0.2865** |
| Up to higher or secondary or further education (A Levels, BTEC etc) | 4 (15%) | 71(14%) |  |
| University or college | 10 (38%) | 222 (52%) |  |
| Post-graduate degree | 12 (46%) | 135 (32%) |  |
| **Employment** |  |  | **p = 0.0523** |
| Employed or self-employed | 22 (85%) | 387 (90%) |  |
| Unemployed | 1 (4%) | 3 (1%) |  |
| Homemaker | 3 (12%) | 34 (8%) |  |
| Student | 0 (0%) | 4 (1%) |  |
| **Last cervical screen (self-reported)** |  |  | **p = 0.0059** |
| Never | 1 (4%) | 18 (4%) |  |
| Over 3 years ago | 10 (38%) | 63 (15%) |  |
| Within the past 3 years | 15 (58%) | 347 (81%) |  |
| **Ethnic group** |  |  | **p = 0.8069** |
| White (any nationality) | 25 (96%) | 407 (95%) |  |
| Any other ethnic group | 1 (4%) | 21 (5%) |  |

**Table 1. Demographic data**
| Birth of last child |  |  | p<0.0001 |
| --- | --- | --- | --- |
| Currently pregnant | 2 (8%) | 52 (12%) |  |
| Within the past 12 months | 20 (77%) | 132 (31%) |  |
| 1-2 years ago | 0 (0%) | 87 (20%) |  |
| 2-3 years ago | 3 (12%) | 77 (18%) |  |
| ≥3 years ago | 1 (4%) | 80 (19%) |  |

**Table 2.** Participant characteristics Number of participants = 26. E, W, S, NI or Bri = White English, Welsh, Scottish, Northern Irish or British origin; m = months; yrs = years.

| Participant number | Screening history | When did give birth (how many children) | Willing to take part in study cervical screen 6weeks? | Willing to take part in study cervical screen if urine sample 6 weeks | More like to have smear after pregnancy if at 6 week check | More likely to have cervical screening 6weeks if only urine sample | Would prefer to do self-testing with urine sample than smear | Prefer cervical screening 6 weeks than 12 weeks | Ethnicity | Age | Employment | Highest education |
| --- | --- | --- | --- | --- | --- | --- | --- | --- | --- | --- | --- | --- |
| 115 | Within last 3 yrs | Within last 4m (1 child) | No | Yes | Disagree | Strongly agree | Strongly agree | Strongly disagree | E, W, S, NI or Bri | 24-29 | Employed/self employed | Post-graduate |
| 289 | Within last 3 yrs | Within last 4m (1 child) | Yes | Yes | Strongly agree | Strongly agree | Strongly agree | Neither | E, W, S, NI or Bri | 30-34 | Employed/self employed | College/University |
| 281 | Within last 3 yrs | Within last 4m (2 children) | Yes | Yes | Strongly agree | Strongly agree | Neither | Strongly agree | E, W, S, NI or Bri | 24-29 | Employed/self-employed | Post-graduate |
| 302 | Over 3 yrs | Pregnant (1 child) | Yes | Yes | Agree | Neither | Agree | Neither | E, W, S, NI or Bri | 35-39 | Employed/self-employed | College/University |
| 153 | Within last 3 yrs | Pregnant (0 child) | Yes | Yes | Agree | Strongly agree | Agree | Agree | E, W, S, NI or Bri | 30-34 | Employed/self-employed | Post-graduate |
| 240 | Within last 3 yrs | Within last 4m (1 child) | Yes | Yes | Strongly agree | Neither | Disagree | Strongly agree | E, W, S, NI or Bri | 30-34 | Employed/self-employed | Post-graduate |
| 216 | Over 3 yrs | Pregnant (1 child) | No | Yes | Neither | Strongly agree | Strongly agree | Neither | E, W, S, NI or Bri | 40-44 | Homemaker | College/University |
| 241 | Within last 3 yrs | 4-12m ago (2 children) | Yes | Yes | Strongly agree | Strongly agree | Neither | Strongly agree | E, W, S, NI or Bri | 24-29 | Employed/self-employed | College/University |
| 016 | Over 3yrs | Within last 4m (1 child) | Yes | Yes | Disagree | Strongly agree | Strongly agree | Disagree | E, W, S, NI or Bri | 24-29 | Employed/self-employed | College/University |
| 030 | Within last 3 yrs | Within last 4m (2 children) | Yes | Yes | Neither | Neither | Strongly disagree | Strongly agree | E, W, S, NI or Bri | 30-34 | Employed/self-employed | Higher/secondary/further education (A Levels) |
| 120 | Within last 3 yrs | Within last 4m (1 child) | Yes | Yes | Agree | Neither | Disagree | Agree | E, W, S, NI or Bri | 24-29 | Employed/self-employed | Post-graduate |
| 186 | Within last 3 yrs | 4-12m ago (3 children) | Yes | Yes | Neither | Neither | Disagree | Neither | E, W, S, NI or Bri | 30-34 | Employed/self-employed | College/University |
| 212 | Within last 3 yrs | 2-3 yrs ago (1 child) | No | Yes | Neither | Agree | Agree | Neither | E, W, S, NI or Bri | 24-29 | Employed/self-employed | Post-graduate |
| 362 | Within last 3 yrs | 4-12m ago (1 child) | Yes | Yes | Strongly agree | Strongly agree | Strongly agree | Agree | E, W, S, NI or Bri | 30-34 | Employed/self-employed | College/University |
| 025 | Over 3 yrs | 3-4yrs ago (1 child) | Yes | Yes | Agree | Disagree | Disagree | Neither | E, W, S, NI or Bri | 35-39 | Employed/self-employed | College/University |
| 202 | Within last 3 yrs | Within last 4m (1 child) | No | Yes | Strongly disagree | Strongly disagree | Strongly agree | Strongly disagree | E, W, S, NI or Bri | 24-29 | Employed/self-employed | Post-graduate |
| 045 | Over 3 yrs | Within last 4m (1 child) | Yes | Yes | Strongly agree | Strongly agree | Neither | Agree | E, W, S, NI or Bri | 24-29 | Employed/self-employed | College/University |
| 348 | Over 3 yrs | Within last 4m (3 children) | Yes | Yes | Strongly agree | Strongly agree | Strongly agree | Strongly agree | E, W, S, NI or Bri | 35-39 | Unemployed | Higher/secondary/further education (A Levels) |
| 093 | Over 3 yrs | 4-12m ago (2 children) | Yes | Yes | Strongly agree | Neither | Strongly disagree | Neither | E, W, S, NI or Bri | 30-34 | Homemaker | College/university |
| 185 | Within past 3yrs | Within last 4m (3 children) | Yes | Yes | Strongly agree | Agree | Neither | Strongly agree | E, W, S, NI or Bri | 30-34 | Employed/self-employed | Higher/secondary/further education (A Levels) |
| 365 | Over 3yrs | 4-12m ago (1 child) | Yes | Yes | Disagree | Disagree | Agree | Agree | Any other ethnic group | 30-34 | Employed/self-employed | Post-graduate |
| 049 | Within past 3yrs | 2-3yrs ago (1 child) | Yes | Yes | Neither | Neither | Agree | Neither | E, W, S, NI or Bri | 30-34 | Employed/self-employed | Post-graduate |
| 051 | Over 3yrs | Within past 4m (3 children) | Yes | Yes | Strongly agree | Strongly agree | Strongly agree | Strongly agree | E, W, S, NI or Bri | 35-39 | Employed/self-employed | Post-graduate |
| 071 | Never (24) | Within past 4m (2 children) | Yes | Yes | Strongly agree | Strongly agree | Strongly agree | Agree | E, W, S, NI or Bri | 24-29 | Homemaker | Higher/secondary/further education (A Levels) |
| 161 | Within past 3yrs | Within past 4m (2 children) | Yes | Yes | Strongly agree | Agree | Agree | Strongly agree | E, W, S, NI or Bri | 30-34 | Employed/self-employed | Post-graduate |
| 385 | Over 3yrs | Within past 4m (1 child) | Yes | Yes | Disagree | Strongly agree | Strongly agree | Disagree | E, W, S, NI or Bri | 24-29 | Employed/self-employed | Post-graduate |

Compared to non-interviewees, participants were no more likely statistically to be willing to be part of a clinical trial of conventional cervical screening 6-weeks after delivery (interview participants = 22/26 (85%) versus non-participants = 286/428 (67%); χ^2^ (1) = 3.557; p = 0.059). Interview participants were, however, more likely to be willing to take part in a study if this involved just urine self-sampling at 6-weeks postnatal (26/26 (100%) versus non-participants 330/428 (77%); χ^2^ (1) = 7.59; p = 0.0059). Interview participants were as likely to think that offering cervical screening at the 6-week postnatal check-up would increase their likelihood of taking up cervical screening (χ^2^ (4) = 2.81; p = 0.059) Whilst some of the differences did not reach statistical significance, this may be due to the small sample size and a type 2 error, so the views of the interview participants should therefore be considered within this context.

Three themes were generated from the data: 1) a window of opportunity; 2) am I ready yet? (due to postpartum recovery); and 3) neglect of women’s health (see Table 3). Illustrative quotes are presented with a number.

**Table 3.** Themes and subthemes generated from analysis of verbatim transcripts.

| Theme | Subtheme |
| --- | --- |
| 1. A window of opportunity | 1.1. It's not just about me anymore |
|  | 1.2. Fitting in screening |
| 2. Am I ready yet? Postpartum recovery | - |
| 3. Neglect of women's health | 3.1. The need for consistent and accurate information |
|  | 3.2. Inadequacy of maternal focus during postnatal care |

### Theme 1: A window of opportunity

Women considered cervical screening within the context of parenthood and the additional importance of maintaining good health that this creates, alongside whether the postnatal period is a suitable time to personally participate in the NHS CSP.

### 1.1 It’s not just about me any more

It was clear that attending cervical screening was viewed as an important aspect of women’s health across the participants’ accounts, especially where they personally had or knew of others requiring subsequent further tests or treatment. Participants described an additional shift in this mentality when thinking about their children. Rather than attendance at screening being an individual decision, having had a baby, participants described their renewed responsibility as a parent to participate in screening.

> *‘Because sometimes you don’t have any symptoms at all, and that’s why it is so important. And the longer you leave it to find out, the more chance that it’s not curable. And especially with having a family and having children, I think it’s just really, really important to go for them.’ (Pt241)*

> *‘I would just rather know…I think after having children as well you feel…your mind slightly changes in terms of you want to be the best version of yourself you can be, and you’re in the mindset of being like I’ve got to be around for my children.’ (Pt 281)*Even in the case of the single participant who had never previously engaged with cervical screening, emphasis was placed on the obligation to be able to fulfil the role of motherhood. Consequently, HPV self-sampling was viewed positively by allowing the opportunity to enact this whilst already being a familiar aspect of interacting with healthcare during pregnancy given that completion of urine sampling is frequently required during this time.

> *‘…making sure that I’m healthy enough to look after them […] I think the urine test would be my first preference. I mean, you do loads of them while you’re pregnant and just after anyway.’ (Pt 216)*

### 1.2 Fitting in screening

Many participants reflected on the challenges of fitting cervical screening into busy daily life and how booking systems used in primary care can make it difficult to schedule their appointments. Resultingly, when weighing up whether screening should be offered additionally during postnatal checks, the idea of screening taking place at a time when they would already be at the same location where it typically takes place made sense. Participants viewed this as an opportunity to capitalise on when they would already be engaged with healthcare services.

> *‘’Cause also, with a new baby, you know, that’s the last thing on your mind, you’ve got a million and one things to think about so if you ask them when they’re already there, it’s kind of two birds, one stone type thing.’ (Pt202)*This especially resonated for those with multiple children, where the practical logistics of managing to attend a specific screening appointment can be particularly burdensome, and for those concerned about whether they could tolerate the screening test procedure should their newborn be present. The uptake of such an offer however could hinge on having sufficient wider social support.

> *‘If you’re already there, chances are you’ve probably already arranged childcare for any other children you have, or your partner might be able to get that day oL to help. So, yeah, having it done there or I don’t know if you can do it as early as when you’re in hospital after having the baby, because you’ve got to be checked anyway, that would probably make it easier.’ (Pt281)*Some participants therefore considered self-sampling as something they could complete at home and a practical solution to ‘fit in’ screening during an unpredictable time-period which could afford them the opportunity to engage in screening on their own terms. This was, however, caveated with the risk that one might forget to complete self-sampling and therefore should not be the only option during this period.

> *‘I think…pretty obviously, being at home and having to do the tests at home would be so much easier than going out, particularly at six weeks, leaving the house feels huge […] being at home would be 100 per cent easier.’ (Pt289)*Overall, these potential adaptations to the healthcare system resonated with participants framing it as a more flexible, woman-centred approach that acknowledges the additional demands on their time and capacity particularly in early motherhood.

### Theme 2: Am I ready yet? Postpartum recovery

Women reflected on how they negotiated or would negotiate recovery from childbirth whilst considering how cervical screening might be integrated into this period. This tied in with readiness to be exposed to a speculum-based examination that requires vulnerability and bodily exposure.

For many participants, the idea of undergoing cervical screening shortly after birth brought a renewed focus to the areas of their body that could be still healing during that time. Their accounts were multi-faceted encompassing emotional, psychological and relational dimensions in addition to physical recovery. Screening was described as potentially too intrusive and possibly re-traumatising particularly whilst participants recalled their postnatal physical recovery experiences.

> *‘I think the thought of having any kind of vaginal examination at the time would have just made me really anxious and upset. Even though I’m sure physically when things have healed it would have been fine, it’s more, like, the mental side of things with the anxiety from that experience.’ (Pt153)*

> *‘So the last thing you want to do is, six to eight weeks after giving birth, and some people have had it really rough and have had really traumatic births, to have someone stick something up there when you’re already feeling possibly emotionally or physically not prepared. So I think it’s just a tricky one. For me, I definitely would have not gone for it, despite me having a very straightforward labour, without any trauma.’ (Pt365)*Concerns about bodily presentation were expressed in relation to embarrassment and fear about exposing oneself or being touched during a screening procedure that could potentially cause pain and whilst still experiencing postpartum bleeding. Additionally, having the mental bandwidth to appear presentable whilst navigating early motherhood was perceived as a necessary pre-requisite in some cases. These views appeared rooted in a desire to preserve dignity and control about when they could ‘be seen’ by others.

> *‘I wouldn’t be that keen on it at the same time. And like, it does seem quite soon after, just in terms of…this is going to sound disgusting, but in terms of actually being clean and dressed properly and all that kind of thing. Because everything goes out of the window when you’re a new mum and you have a new baby’ (Pt161)*This was additionally interwoven in several accounts related to sexual identity and intimacy where a return to sexual activity was viewed as a milestone or marker of sufficient healing; only after this timepoint, could participating in cervical screening be considered.

> *‘I think if the first-time post-birth was going to be a smear, I don’t know. That makes it feel daunting. Knowing that I’ve been intimate, and that I can handle that, would make me think I could handle doing a smear test.’ (Pt 240)*However, perspectives about this time-period varied with some participants highlighting that the frequency of intimate examinations during pregnancy and childbirth appeared to normalise how they viewed procedures such as screening. For these women, screening appeared less likely to provoke discomfort or resistance when perceived as part of bodily checks conducted by others. Similarly, successfully completing speculum-based screening, and therefore being examined to receive feedback on healing, could provide some participants with confidence that they would be able to resume sexual activity.

> *‘I had [baby’s name] and having a vaginal delivery, having…you know, kind of got used to being prodded and poked and stuL so many times medically as well as men sort of more privately […] And then it was just a lot…I wasn’t as nervous because I was just like, yeah, this is not the worst thing that’s ever happened to me. And then it was also the discomfort wasn’t really there. And I guess that’s something to do with having birthed a whole human through there a speculum didn’t feel quite so uncomfortable.’ (Pt212)*

### Theme 3: Neglect of women’s health in and around pregnancy

Participant’s views on cervical screening and self-sampling were shaped by wider views on experiences of support and levels of how informed they felt about their healthcare during pregnancy and postnatal periods. Postnatal care for themselves in addition to their baby was, for some, described as incomplete or tokenistic and so cervical screening could become another area where women perceive their health needs are sidelined, including if the screening test were to change to self-sampling only. These views were also influenced by our previous exposure to the free text comments in the questionnaire study [28].

### 3.1 The need for consistent and accurate information

Participants repeatedly emphasised the importance of clear, timely, and consistent information about cervical screening during the pregnancy and postnatal period. Women highlighted receiving mixed messages and unclear advice creating unnecessary confusion and annoyance.

This included receiving screening invites whilst pregnant then being told they would have to wait until postpartum with health professionals often unable to confirm when screening could take place post-birth.

> *‘… it was also, like, frustrating for the person making the calls and sending the letters because it seems like a bit of a waste of time and money. But it seems like a bit of a system failure that no one’s really worked out a cure for yet and it will just persist because it’s not a priority.’ (Pt025)*Some therefore questioned whether the potential to offer cervical screening during postnatal checks is due to a heightened risk of cancer at that time leading to some unease and reflecting an uncertainty about the intentions behind this proposal. If left unresolved, inconsistent information could risk causing women feeling left to navigate their own care during an already overwhelming time and might ultimately distrust the offer.

> *‘I feel like I quite want to go for a test, I feel like I’ve been for so long now without having a smear and there has been lots of changes to my body and I do wish that they called me up and I do feel a bit unsure about where I stand with it. I don’t know how long after, nobody said to me, because when you go for a smear, they always say, are you pregnant but then throughout pregnancy, they don’t say to you, oh when you’ve had your baby, it’s going to be this long until you have your smear.’ (Pt120)*Equally, despite initial positive reflections around the concept of self-sampling as being akin to familiar types of tests, such as those used in sexual health, discussing self-sampling testing may exacerbate the feeling of receiving inadequate communication from healthcare services. Participants therefore described a desire to receive easy-to-follow instructions and sufficient reassurance about self-sampling test accuracy compared with speculum-based screening tests to alleviate apprehension.

> *‘I suppose, it depends on…is it…would the urine and the self-done swabs be as accurate as actually having it done at the doctors? I suppose that would depend on it for me.’ (Pt185)*

> *‘Well my immediate thought is how, in terms of them done by a professional as opposed to one’s self, what is the sensitivity of those tests compared like…because they’re, yeah, a big diLerence in sensitivity […] I guess the worry of not doing it properly, of getting false negatives. Mostly, yeah, the idea that it may not be as accurate if it’s done oneself, unless the instructions are really clear and precise*.’ (Pt365)

### 3.3 Inadequacy of maternal focus during postnatal care

Participants shared that they felt their own health was deprioritised after birth, compared to their baby’s. Some highlighted that postnatal appointments did not meet their emotional and physical needs. This perceived ‘slipping through the net’ influenced how they viewed the introduction of cervical screening during this time. On the other hand, speculum-based cervical screening was sometimes viewed as increasing the likelihood of receiving a more comprehensive mother-focused appointment with the potential to addressing wellbeing concerns experienced by mothers that could otherwise go unnoticed.

> *‘I’d quite welcome having some kind of examination just to make sure that everything’s going back to normal. And then part of that would be a smear test, and that would be fine for me. […] just like a full, kind of, little MOT, post birth, would be quite nice. Yeah, I’ve never had that so I suppose part of having that done at the same time as the smear would just give you opportunity just…like, if you’d had an episiotomy or a tear or anything like that, just, kind of, check that at the same time.’ (Pt186)/*

> *‘…encouraging women to go before 12 weeks or 16 weeks or whatever, not that long after giving birth would have given people the opportunity to be checked down there, because we’re kind of left to it.’ (Pt115)*Careful consideration of how this offer, and the inclusion of self-sampling, is communicated is important to minimise exacerbation of negative perceptions views to ensure it is conveyed as beneficial to ‘the patient’, not just the healthcare system. Choice was therefore highlighted as an important factor to be integrated into any proposed change.

> *‘it’s not something that I would rule out. But what I would be really keen to see is that if you choose not have it at six weeks, it’s still fine to have it later on down the line, that you’re not pushed to have it done too soon. Because I know obviously there are shortages of staL and they want to get appointment doubled up rather than having you come in separately.’ (Pt.016)*This underscores how willingness to engage with cervical screening was not only shaped by whether the opportunity is convenient or participant’s individual readiness but by whether healthcare interactions are experienced as supportive of the woman.

## Discussion

To our knowledge, this is the first study to use qualitative interview methods to explore what women think about the offer of cervical screening during the postnatal check-up.

### Summary of findings

Women recognised the importance of cervical screening but recognised barriers to attending screening. As new parents, they were busy, struggled to get out of the house in a ‘presentable’ state and prioritised care of their babies. Nonetheless, there was a recognition that it was now more important for them to look after themselves, as they had parental responsibilities.

Practical barriers, including primary care booking systems, added logistical barriers to views about screening and many felt that offering screening at the postnatal check-up would make these aspects easier. However, there was some concern that people may not be physically or psychologically ready for a speculum examination at the postnatal check-up. Urine self-sampling was an attractive option because of this, although there were concerns about accuracy and remembering to complete and return it, if sent in the post. However, others felt that, in comparison to childbirth, a speculum examination was much easier and welcomed the opportunity to have someone check that their vagina and perineum had healed, giving reassurance about returning to sexual activity. Frustration around receiving mixed messaging and disinformation around cervical screening in and around pregnancy was highlighted and there was a sense that women felt deprioritised compared to their child’s health, and a shift in focus to maternal care aspects, would therefore be welcomed.

### Strengths and limitations

A strength of this study is that it arose out of a larger questionnaire with 454 participants and the results built on themes that arose from analysis of this much wider sampling. This study was informed by these earlier data and gave us the opportunity to explore ideas generated from analysis of free-text responses in greater depth, checking our understanding of brief comments had not been misinterpreted. The topic guide was therefore informed by participant responses from this broader, but shallower, information base. Free-text questionnaire comments from participants of the earlier questionnaire study were not used to illustrate themes from the earlier study, so the ideas expressed here represent new views [28]. Because of the wide geographical sharing of the original questionnaire, via social media and clinical research networks across England, these responses are not specific to a particular GP practice cohort or obstetric unit, so are likely widely applicable.

A significant limitation is that there were few participants who had never engaged with cervical screening, although 10/26 were out of date for cervical screening, in line with our previous data [8]. Another limitation is that participants had relatively high levels of educational attainment and the majority were employed, or had been prior to maternity leave, and those from minority ethnic groups not well-represented (1/26). This is despite sharing the questionnaire widely, including maternity groups from inner city areas of Bristol and Manchester, and having participants in the questionnaire study likely to be from higher Index of Multiple Deprivation (IMD) backgrounds. Participants from other backgrounds did initially offer to be involved but did not, or did not feel able to, respond to email invitation. This perhaps reflects less confidence to engage with research generally, or that their views would be considered, and could be contributed to by lack of access to electronic devices, lower levels of literacy, lack of English as a first language, lack of childcare to facilitate a prolonged conversation etc. Telephone consultations were offered, which may have on-set this barrier, as would the offer of remuneration for time to conduct interviews. It would be interesting to repeat these conversations with direct approach to mothers via community groups and via in-person contact at the time of the postnatal check-up. This is something we will explore in further work using differing recruitment strategies and supported by in-person translators, to directly target populations more likely to be under-screened. We will also explore this in an ongoing clinical study of paired sampling at 6- and 12-weeks postnatal, in a sub-study looking at demographics of consecutive women approached for recruitment and compare whether there is a difference in demographics of those who choose to participate and those who decline when approached in person, during pregnancy and after delivery. Interestingly, there are regional and racial disparities in those who attend the postnatal check-up, reflecting lower general engagement with healthcare and research in some groups, which we need to try harder to reverse [33].

### Relevance to existing literature

Previous literature has identified that primary care practices in England have insufficient time to conduct ‘gold standard’ postnatal consultations [34].Our findings add to the growing evidence that those offered self-sampling are positive about the idea but must be provided with sufficient information around the accuracy of these compared to speculum-based screening tests and that there is variation in personal preferences [10,19,35]. As such, it has been suggested that multiple test types could be offered in practice although women may ultimately want to receive a recommendation about which to choose. [36]

The way participants felt their sense of self had changed after childbirth influenced the importance they attributed to cervical screening. However, whether they saw it as important or not, this change in self-identity still affected how able they felt to attend screening at the postnatal check. The postnatal period has previously been identified as significant in how one views the self and how it can be altered [37].

### Implications for future research and practice

There is a need for communication strategy to be developed for women in and around pregnancy and for those who care for them, similar to a recent project focusing on decision-making about cervical screening test type [38]. Whilst participants in this study deemed cancer early detection important, the need expressed by some for improved maternal postpartum care at the system-level risks introduction of self-sampling being interpreted as further withdrawal of the healthcare system’s duty of care towards women. Any future shift in practice requires careful messaging and support for other opportunities for in-person care.

The findings from the on-going PINCS research programme will provide evidence of acceptability and feasibility of both a paired-sample study and a randomised control trial of cervical screening at 6 and/or 12 weeks postnatal. The results of these ongoing and future studies could influence adaptations to what is currently considered essential for postnatal care [39].

Future research should also explore the views of primary care health professionals involved in pregnancy and postnatal care consultations, as health-related behaviour change conversations are underutilised in this setting and primary care physicians, health visitors, practice nurses and midwives may not view offering cervical screening to be part of these appointments [40].

## Conclusion

Offering cervical screening alongside the postnatal check-up would reduce some of the practical barriers to cervical screening in postnatal women. However, this may be too soon for some, especially those with physically and/or emotionally traumatic obstetric experiences. This might also give opportunities to identify those requiring more support and a greater focus on women’s health issues after childbirth would be welcomed. These data support the need for further studies to explore the value of earlier cervical screening in the postnatal period, as this may improve uptake. Further work, specifically targeted on harder to reach groups would be of value.

## Data availability statement

The data that support the findings of this study are available on request from the corresponding author. The data are not publicly available due to privacy or ethical restrictions.

## Ethics approval statement

The study was approved by Hampshire A Research Ethics Committee on 30/3/23 - IRAS project ID: 321358; REC reference: 23/SC/0082; 12/1/24 amendment reference: 20240105.

## Participant consent statement

Individual participant consent was obtained.

## Permission to reproduce material from other sources

Not applicable

## Clinical trial registration

Trial was registered with the National Institute for Health and Care Research Central Portfolio Management System (CPMS ID: 55489) and https://bepartofresearch.nihr.ac.uk/

## Author Contributions

Conceptualisation - JM with the aid of LMcW, EC and SS.

Methodological design - JM and LMcW with contributions from RN, HBR,EC and SS. Funding acquisition – JM with substantial contributions from LMcW, SS and EC.

Study materials for ethical approval were designed by HBR, RN, JM, LMcW with input from EC and SS.

Analysis – RN, LMcW and JM.

Investigation - interviews conducted by RN. Resources - JM.

Writing - Original Draft – VC, JM, and LMcW Writing - Review & Editing – all authors Validation –LMcW and JM.

Visualization – JM and LMcW Supervision – JM and LMcW Project administration – JM

The final version was approved by all authors. JM acts as study guarantor.

## Data Availability

All data produced in the present study are available upon reasonable request to the authors

## Acknowledgements

We are grateful to Flo Cullen, Karen Tanner and Richard Innes of the Department of Research and Development at Somerset Foundation Trust for their substantial enthusiasm and support in setting up this study. We thank GO Girls, Jo’s Cervical Cancer Trust (sadly no more), Maternity Voices Groups, especially those in Somerset, Mumsnet and the British Gynaecological Cancer Society for their feedback and support to advertise this study, and the members of the NHS Cervical Screening Programme Research Innovation, and Development Advisory Committee for their invaluable advice and support. Finally, we thank the participants in pre-PINCS for sharing their thoughts and ideas about this and future studies and those in our previous focus groups; this work was at the request of new mothers, and it is our honour to work towards putting your ideas into practice.

