## Supplementary materials 1 for "A qualitative study investigating the acceptability of cervical screening and self-sampling in postnatal women at 6-week postnatal check-up"

**Topic Guide: Pre-PINCS semi structured interviews**

Interview objective

To further understand attitudes to postnatal cervical screening tests and participation in associated studies

Qualitative project aims

1. Assessment of acceptability of, & attitudes towards, liquid based cytology & HPV screening at 6- vs. 12-weeks postnatal in pregnant & recently pregnant women.
2. Feasibility and acceptability of self-testing for HPV using urine tests at 6- & 12-week postnatal compared with conventional LBC testing at 6- & 12-weeks

Methods

Purposive sampling will be used to invite those who indicate willingness to participate in an interview based on recency of pregnancy, cervical screening status (e.g., never attender) & other demographic factors collected during completion of the survey. If there are insufficient volunteers from the survey, we will invite women who have delivered through our services.

Approximately 30 women will take part in an individual semi-structured interview.

A topic guide will be developed, informed by the Theoretical Framework of Acceptability (Sekhon 2017, Sekhon 2021). Interviews will be audio-recorded with consent, transcribed verbatim & anonymised. The anonymised transcripts will be analysed using a form of Thematic Analysis (Gale, 2013)

Sekhon M, et al. Acceptability of healthcare interventions: an overview of reviews and development of a theoretical framework. BMC Health Services Research. 2017;17(1):88.

Sekhon M, et al. Pregnant and breastfeeding women's prospective acceptability of two biomedical HIV prevention approaches in Sub Saharan Africa: A multisite qualitative analysis using the Theoretical Framework of Acceptability. PLoS One. 2021;16(11):e0259779.

Gale NK, et al. Using the framework method for the analysis of qualitative data in multi-disciplinary health research. BMC Medical Research Methodology. 2013;13(1):117.

This topic guide has been designed to explore the attitudes to cervical screening and more specifically to understand views on the timing and methods of cervical screening in the postnatal period.

**Interview**

**Introduction**

**Start recording**

Thank you for completing our online questionnaire and agreeing to participate in this online interview. Before we start, I’d like to remind you about our study. We are gathering information about what people think of cervical smear tests after having a baby. I would like to know what you think about the different ways this test can be performed and when might be a good time for the tests to be offered. I want to know how you would feel about taking part in cervical screening studies in the weeks and months after birth.

There are no right or wrong answers to my questions. It is completely up to you whether you decide to take part today or not. If you decide you don’t want to take part anymore, just let me know. You can stop the interview at any point.

If you don’t want to answer any of the questions that is fine, just let me know and we can move on to the next question. It might sound like I am repeating myself, but I am just checking I have understood what you have said. If you don’t understand some of my questions, just say and I will try to rephrase them.

If it is OK with you, I will record the interview. Afterwards, we will type up what you said, but anything that could identify you, such as your name, will be taken out. No one outside of the research team will know that you have taken part in the study.

I have planned a few questions and things for us to talk about, but if you have anything else you want to say or add at any point, please feel free to do so.

**Confirmation of identity**

**Confirmation of consent**

1.	I confirm that I have read the information sheet (Version 1.1 dated December 2023) for the above study. I have had the opportunity to ask questions and understand why the research is being done.

2.	I understand that my participation is voluntary and that I am free to withdraw at any time by stopping the interview or by contacting any of the individuals above without giving any reason and without my medical care or legal rights being affected.

3.	I understand that the audio from my interview will be recorded and transcribed. This data will be stored securely and anonymously as approved by the Research Ethics Committee. I consent for these to be stored for no more than 10 years.

**Stop recording**

**Re-start recording**

I understand from the survey that you already have some thoughts on/experience of cervical screening so I wonder we if could start with that

- What I’d like to talk about today relates to the answers you gave to the questions in the survey about cervical screening. You don’t have to remember your answers as I have a record of them and you might remember something else or change your mind and that’s okay
- *a little bit tricky to ask knowledge based questions first so suggest starting with cervical screening experience in general – they will say if they aren’t quite sure about what it is etc*.
- From your answers in the survey you completed, you told us you have [never been to cervical screening/you haven’t been to screening in the past 3 years/you have been to screening within past 3 years]. I’d like to start from there and can you tell me more about this?
  - What made you decide to go/not go?
  - What made it easy?
  - What made it difficult? *PROBE: childcare, timing of appointments, availability of appointments, booking system, travel*
  - How do you feel about going to cervical screening in the future? (Why)
  - Is there anything that could make it easier to/you more likely to attend in the future? (Why)
- We know that some new mothers and staff in GP practices think it might help women to have cervical screening if they are **offered** it at their 6-week post-natal check-up, what are you first thoughts about this?
  - What is good about this idea? *PROBE: appointment already booked, contraception, reassurance from examination*
  - What is bad about this idea? *PROBE: birth and healing, bleeding, pain, trauma, worry about results, how and when do you feel would be the right time to offer cervical screening*
- You answered that you had heard of HPV, remember this is not a test but can you tell me what your understanding of it is?
  - Can you tell me about how it relates to cervical cancer and the cervical screening programme we have?
- There is on-going research looking at different ways of testing for HPV, have you heard about any of this or taken part?
  - If yes, ask to elaborate
  - If no, explain that there are self-taken tests like the coli-pee and self-taken swab *(here it might be helpful to have visual aid comparing current screening with the two alternatives, explain the coli-pee and self-swabs only detect HPV and so a smear test would need to be taken if HPV was detected)*
- What are your thoughts on [collipee / self-taken swab / any other formats ] instead of current cervical screening?
  - How do you feel about being offered it?
  - How do you feel about using it?
  - What do you think are positives about offering this?
  - What do you think are negatives?
  - How confident would be in using it? *PROBE: we don’t yet know how well these different types of tests work in women who have recently had a baby. Would it be more important to you to have a test that is non-invasive or the test that is most likely to work or be most accurate?)*
- You mentioned in the survey that you [would be/would not be] willing to take part in a study that involves having a cervical smear 6 and 12 weeks after delivery, can you tell me more about why you said [yes/no]?
- You mentioned in the survey that you [would be/would not be] willing to take part in a study that involves providing a urine sample 6 and 12 weeks after delivery, can you tell me more about why you said [yes/no]?
- Would you be more inclined to participate if:
  - There was only one visit
  - We used self-swabbing rather than urine samples
  - You did not have to attend the hospital/the samples were performed at home/at the GP practice
- Can you tell me about any ways you think would be effective in promoting participation in studies?
- I’ve now covered everything I hoped to talk to you about but do you have anything to add? Is there anything you thought we would talk about and we haven’t that you’d like to now?

**Stop recording**

Thank you again for taking part in this interview. This recording will now be typed up but all identifiable information will be removed or replaced. If you have any questions after today then please do not hesitate to contact me or the team - the contact information is on the patient information sheet we sent to you.
